# A survey approach to assess the work and learning environment and stimulate informed change within the individual departments of an academic health center

**DOI:** 10.64898/2026.01.23.26344679

**Authors:** Sapna Varkey, E. Nathan Thomas, Rebekah L. Layton, Ashalla Magee Freeman, Snigdha Sompalli, Rachael Manasseh, Makayla King, Michel D. Branch, Stephanie H. Brown, Donald E. Pathman

**Author notes:** Corresponding Author: (SV). These authors contributed equally to this work (Sapna Varkey, E. Nathan Thomas, Donald E. Pathman). These authors also contributed equally to this work (Rebekah L. Layton, Stephanie H. Brown). ENT and DEP are Joint Senior Authors.

## Abstract

Creating a psychologically safe and supportive environment is important for everyone who works and learns within an academic health center (AHC) as well as important to AHC overall success. Recognizing a need to better understand the strengths and areas for improvement in an AHC’s environment, we developed and fielded an environment survey tailored to our many student, resident, postdoctoral researcher, staff, and faculty constituencies. This paper outlines the methodological and practical considerations and outcomes in developing and deploying the survey, including information on how it was received and the activities it subsequently stimulated. Three elements were central to our efforts: (a) analyzing and reporting findings at the individual department, center, and learner group level, where individuals directly experience the environment day-to-day, (b) characterizing the environment through three core dimensions of people’s experiences, which were perceiving that the department strives to be supportive; feeling embraced, appreciated, and supported within the environment; and feeling accepted in one’s beliefs, and (c) prioritizing the dissemination of findings to all members of the community. We provide our questionnaire and outline the findings report design, a document received favorably by departments and their leaders. All of these documents can be adapted by other AHCs aiming to assess their environments, learn where to target interventions, while also monitoring progress.

## Introduction

Like all healthcare and teaching institutions, academic health centers (AHC) are their strongest when learners, staff, and faculty in all positions and of all backgrounds feel psychologically safe (e.g., ability to speak up without fear or punishment) and supported in the places where they work and learn [1–4]. Safe, healthy, and supportive environments foster worker productivity, satisfaction, retention, and recruitment [5–8]. Clinical and research teams perform better when members feel psychologically safe and feel they belong [9–10].

AHC leaders cannot know how employees and learners—students, residents, postdocs, fellows—feel about their work and learning environments without directly asking them. Leaders can perceive the environment to be more inclusive and supportive than rank-and-file employees [11]. AHC leaders need data, along with a simple way to understand and translate the findings, where recommendations are outlined in ways that engage their members (e.g., faculty, staff, learners) and to improve department outcomes.

Various AHC’s have previously described their efforts utilizing climate environment surveys (also known as organization climate surveys) to understand campus culture [12], assess the current state of department and organization cultural competence[13–14], identify occurrences of microaggressions, mistreatment, and discrimination within departments [15], diagnose and benchmark the current environment and leverage opportunities for improvement [16], and as part of strategic planning processes for organizational change [17]. The AHC in this paper similarly sought to survey its members to understand people’s experiences in a broad range of areas and to disseminate findings in ways to inform, engage, challenge, and motivate leaders and rank-and-file members of each organization unit and each learner group to stimulate change. Over a two-year period, a survey team designed and fielded an environment survey that built on prior instruments and disseminated findings through individualized reports and presentations to more than 50 departments, centers, offices and learner groups. An encouraging level of engagement across the institution was attained with the survey itself and then with its findings.

This paper outlines the development of the survey instrument and its vetting through focus groups and pilot testing. It outlines our methods of survey fielding to gain strong participation. We describe how findings were crafted into reports that balanced the many details with a broader view of each department’s environment, all benchmarked against data from other departments. We describe how each department’s report was presented to its assembled members, and characterize how this information was received, the questions and feedback generated, and how the early data-informed corrective initiatives it stimulated. We provide guidance and share lessons learned for other institutions undertaking similar initiatives.

## Background

The timing of our 2021-2022 survey initiative coincided with the worldwide social and economic disruptions of the COVID-19 pandemic, the growth of movements that raised awareness of sexual violence and harassment (e.g., the #MeToo movement), and the attention that and other social justice movements sought to bring to the injustices regularly experienced (e.g., murder of George Floyd and Breonna Taylor and the Black Lives Matter #BLM movement) by groups of various gender, race, sexual orientation, immigrant, religious and political identities. Many institutions, including universities and health care organizations, responded to these issues by instituting policies and dedicating resources to improve psychological safety in the organizational culture to better support employees and learners of all backgrounds and beliefs [18–19].

In 2021, the joint leadership of the AHC and its wider health care organization saw the need, based on the national challenges, for an environment survey. Both parts of the organization had separately undertaken environment surveys a few years earlier and had anticipated there would be follow-up surveys. Financial and personnel resources from both parts of the organization were combined to contract with a survey research firm, design and field the survey, and issue an initial report.

Similar to other institutions, this survey followed Urie Bronfenbrenner’s Ecological Systems Theory (EST) [20], which focuses on the interaction and interdependence of individuals (e.g., learners, faculty members, staff members) with their surrounding environments or systems. There are roles, rules, and norms that make up EST’s five systems, or five successively broader spheres of influence, and these ecological systems together influence individuals’ experiences, development, and behaviors. The narrowest system, the microsystem, is the most proximal setting in which an individual is situated and where they spend most of their time and directly interact (face-to-face) with colleagues, work teams, patients, and others. In the AHC context, the microsystem is each department, education program, clinical and research center, hospital unit, administrative office, and outpatient practice. Accordingly, this survey instrument posed questions about individuals’ experiences as they relate to culture, sense of belonging, and support at the microsystem level. These same questions were then again posed for individuals’ experiences at the mesosystem, or AHC-wide level, where the individual departments, hospital units, and offices of the many microsystems connect. Guided by these systems, the overall goal of this work was to strengthen the working and learning environment within the AHC so all staff, faculty and learners would feel not only welcomed but truly valued, and where no one felt marginalized or mistreated to maximize organization outcomes. The objectives of this survey initiative were to (1) develop a questionnaire that broadly measured the AHC’s working and learning environment in the many ways that it can and should support all personnel, as well as identify issues that challenged specific groups, (2) field this questionnaire in ways that generated broad interest and strong participation, (3) use survey findings to inform and engage all members and leaders of the AHC community and then stimulate initiatives to strengthen the environment, and (4) use the questionnaire again in future iterations of the survey to monitor the environment and document improvement with tangible metrics.

### Implementing the survey

#### AHC survey team

An external survey contractor was engaged to help organize and field the survey. Engaging an external survey group added expertise and insights to the survey team, and we anticipated that an external survey group would reassure participants about confidentiality of their responses to promote survey participation and candid reporting.

Nine individuals from both the AHC and its larger health care organization, together with representatives from the survey contractor formed the AHC survey team that planned and oversaw the climate survey. Team members included the AHC executive leader who oversaw maintaining a healthy environment, faculty and staff survey and subject matter experts and project organizers, as well as data systems staff from the human resources department to prepare student and employee lists for the external survey group. The team kept AHC leadership updated on survey progress to maintain leadership’s investment in the project and to coordinate timing of this survey with the AHC’s other surveys and communications with employees and learners.

The AHC survey team foresaw the importance of providing detailed findings about the organizational environment and presented at the unit level (e.g., department, center, administrative office), individuals’ local environment within which they work and learn every day and about which people are naturally most interested in understanding and improving. A rich literature speaks to the effectiveness of feedback provided to teams, defined as “a distinguishable set of two or more people who interact, dynamically, interdependently, and adaptively to achieve specified, shared, and valued objectives” [21–23]. AHC departments are generally larger than the teams studied in the literature, but individuals within departments share valued goals and their constituent divisions generally work closely together. Departments’ relatively large size for “team” feedback provides advantages when it comes to survey assessments, in that data from a greater number of respondents yields more stable parameter estimates of the environment and also permits within-department subgroup comparisons, for example the experiences of women versus men.

#### Survey instrument

The survey instrument’s sections were as follows:

1. Introductory text explained the importance of a supportive environment, the survey’s purpose and the intended use of the findings, and promised confidentiality. Participants were instructed to respond to questions from the perspective of their primary group identity, which they might feel is their professional, gender, ethnic, religious, or generation group. Key definitions were provided, most importantly on which entities/locations should constitute their “department” and “organization,” so that they would report experiences within the settings expected by the survey team and within which data would be analyzed and reported.
2. In the second section, participants were asked to indicate their level of agreement—on a five-point Likert scale, from strongly disagree to strongly agree—with 21 statements about their experiences and perceptions within their specific unit (department, training program, center, office, etc.) and then again within the AHC overall. The 21 items, including “*Perspectives like mine are included in decision making*,” and “*My accomplishments are recognized similar to others’ accomplishments*,” were adapted from existing banks of questions used in others’ environment assessments of their AHCs and other organizations, supplemented with statements addressing other important issues in the environment the survey team felt were missing from previous instruments [24–26]. All statements were intentionally crafted with inclusive language to reflect the wide variety of AHC members’ roles (e.g., faculty, staff, learners) and were vetted with pilot groups to insure survey applicability across roles.
3. The questionnaire’s third section asked participants about experiences with mistreatment, bullying, sexual harassment, microaggressions, discrimination, and physical violence, and about their comfort reporting these occurrences.
4. A briefer, fourth section asked about participants’ knowledge of the organization’s policies to maintain a safe environment, about their interests in future related learning opportunities, and perceived needed areas of training for faculty, staff, and learners.
5. In a final section, participants reported their role in the organization; generation (Millennial, Generation Z, etc.); level of education; racial-ethnic group identity; religious affiliation; location of upbringing (e.g., rural vs urban; U.S. or international); veteran status; and self-identified disabilities, which was information used in the analyses to compare experiences across groups.

#### Focus groups and pilot test

Six virtual heterogeneous focus groups comprised of faculty, staff, and learners were held in the summer of 2021 via Zoom (Zoom Video Communications, San Jose, CA). During a pilot testing phase, prospective participants provided helpful feedback on the overall structure of the survey instrument and its skip patterns, provided important suggestions on the identification and terminology of various demographic groups, and validated the team’s plans for gaining strong survey participation and for maintaining confidentiality and protecting respondents. Participants also verified that they felt most connected to their departments/units (which confirmed the team’s decision to use this as the grouping unit for the organizational survey) and that they appreciated that the survey queried their experiences within their specific departments/units in addition to asking about their experiences in the organization overall.

A pilot test conducted in December 2021 with a sample of 16 faculty, staff and learners using Qualtrics (Seattle, WA) demonstrated that the instrument and survey processes generally worked for the diverse survey cohort.

#### Identifying the survey target population

A member of the survey team from the medical school’s Human Resources Department, in partnership with the University Registrar’s office, identified eligible survey participants within the AHC, which were all students, residents, clinical fellows, post-doctoral fellows, and fulltime and parttime staff and faculty of the AHC. This target population was then assigned to their departments, administrative offices, centers, learner groups and other units within the AHC, which served as the units for which response rates were calculated and within which responses were grouped in unit-specific reports. Best unit assignments for some individuals were not always obvious, and the survey team set several decision-rules for consistency. For example, survey responses from faculty who worked within a given research center or medical service center (e.g., the cancer research center and dialysis center) were assigned to the department where they held academic appointments (e.g., department of genetics or pediatrics), whereas the faculty directors of these centers were assigned to their center.

The survey plans and instruments were submitted to, and found exempt by, the university Institutional Review Board (IRB), determined to not constitute human subjects’ research. The IRB also approved plans for data protection and for data sharing between the survey contractor, the AHC, and the larger health care organization.

#### Fielding the survey, promoting participation, and response rates

The survey contractor emailed invitations to all members of the AHC, with embedded individualized links to the online survey. Authors did not have access to information that could identify individual participants during or after data collection. Invitations indicated the survey’s purpose, how group findings would be shared and used, and promised anonymity for respondents. The survey was open from February 8, 2022, through February 28, 2022. Survey reminder requests were emailed to nonrespondents weekly, for a total of up to four invitation requests. Outcomes reported in this study are based on an existing report for an unidentifiable department, referenced on March 10, 2025.

A variety of approaches were deployed to promote strong survey participation, as we anticipated that a robust response rate was critical for the survey’s results to be convincing and have impact within and across the organization. Before initial survey invitations went out and then again midway through the survey period, the AHC (dean/CEO) emailed the entire AHC community to highlight the survey’s importance and urge participation. A “survey champion” was designated by the chair within each department/unit and charged with galvanizing interest and building participation in the survey. Each unit also had a survey oversight team that included the champion, department chair or division/center/unit director, and the department’s inclusion/engagement liaison (identified by department chair). Oversight teams reviewed their unit’s participation rate reports generated every three days by the survey contractor and strategized additional communications and other approaches to stimulate participation.

Champions were provided with information on “best practices” to promote survey participation. Survey recruitment efforts by individual departments and offices proved more effective than AHC-wide efforts, with the most effective approach being to dedicate time to complete surveys at departments’ regular gatherings, like monthly faculty or staff meetings.

A total of, 4,046 (56%) members of the AHC community responded to the survey, which the survey team and departments considered strong and survey findings thus sufficiently representative. Participation rates varied across units: 55% for faculty and staff within clinical departments (43-93% across individual departments); 58% within basic science departments (21-70%); 82% within AHC administrative offices (65-88%); 45% among residents; 34% among medical students; and 43% among master’s and doctor students (13-70%).

### Disseminating survey findings

#### Initial AHC-wide dashboard reports

The external survey contractor, with input from the AHC/health care organization survey team, created an interactive on-line dashboard with findings for the AHC as a whole. The dashboard presented AHC-wide mean group response values and percentage breakdowns to selected survey items offered as key indicators of the environment (e.g., I have sense of belonging at [the AHC]). The external survey contractor’s dashboard emphasized a visual, readily understood presentation of findings. The findings were presented by the survey contractor to top leadership of the AHC and to department chairs, who could also access the dashboard online.

The strength of the online dashboard was that it provided an early, high-level view of how people regarded key pieces of the AHC’s environment, but its information did not present a full or coherent picture of the environment, or point to specific needed next steps. Department Chairs generally found the online dashboard cumbersome to manipulate and didn’t have the orientation needed to “drill down” to extract useful information. In the end, the environment dashboard did not garner much attention, likely due to challenges in understanding and synthesizing the data.

#### Department/unit reports

Reports for each department/unit were designed to provide information on both the environment broadly as well as details on the types of challenges people face within the environment and the groups affected. The report format was designed to be appropriate for an initial group presentation to each unit and to also then serve as documentation and benchmarking for aspects of the environment the unit will address going forward.

Key design features of unit-level reports were:

- *Condensing data into a few key dimensions of the environment.* Respondents’ agreement or disagreement with the 21 statements about their experiences and perceptions of the environment within their units were collapsed down to fewer underlying dimensions that could be more easily communicated, understood and remembered. Exploratory factor analysis identified 16 statements that could be reduced to cleanly load onto their three underlying dimensions of the perceived environment: (1) perceiving that the unit strives to be supportive, (2) feeling embraced, appreciated, and supported within the environment; and (3) feeling accepted in one’s beliefs. (Table 1)
- *Presenting comparison group data.* No matter one’s numeracy, it means little to learn that their department’s average response value on a 1 to 5 scale of, say, feeling embraced, appreciated, and supported within the environment, is a 3.5, a mid-value between “neither agree nor disagree” and “agree.” It generally means more to learn that the average response within one’s department on a 1 to 5 scale of feeling embraced, appreciated, and supported within the environment scale is 0.5 units lower than that of other clinical departments within the AHC which placed it in the bottom quartile of all departments (Fig 1). Displaying anonymized comparison data for other, related units on the various environment metrics highlights where a department exceeds and where it falls short of peer units, which naturally conveys a sense of what one’s department could and should achieve. Comparative data also naturally suggests the targets for any subsequent interventions, e.g., “Our goal is to be in the top quartile of departments in our members feeling accepted in their beliefs.” Comparison groups used in reports were, as appropriate, all other clinical departments, all other basic science departments, other divisions within a given large department, and other administrative offices.
- *Stressing the magnitude of between-group differences rather than the statistical significance of differences.* The purpose of presenting comparison group data was not to identify measures on which there was a statistically significant difference in group (e.g., department) means or proportions, that is if there was less than 1 in 20 chance that a difference of that size occurred by chance alone. Rather, a greater difference found in mean responses on an environment measure for one unit compared to other units, or between subgroups within large units, was intended to more broadly signal that the difference was more likely real and, more importantly, more likely a meaningful difference in the experiences of the people involved. This does not require inferential statistics. And by not including *p*-values or confidence intervals for the many metrics presented with comparison data, reports were visually less cluttered and more easily understood by all individuals. Summaries of key findings within the reports and in presentations to departments emphasized findings where group response mean and proportion differences were greatest and, given respondent group sizes and unit response rates, were least likely to be due to chance alone.
- *Protecting respondent anonymity*. In presenting comparison data for subgroups within departments, no data, tables or graphs were presented for any group with fewer than five respondents. Accordingly, reports for larger departments included more subgroup comparisons than reports for smaller units.

**Figure 1.**
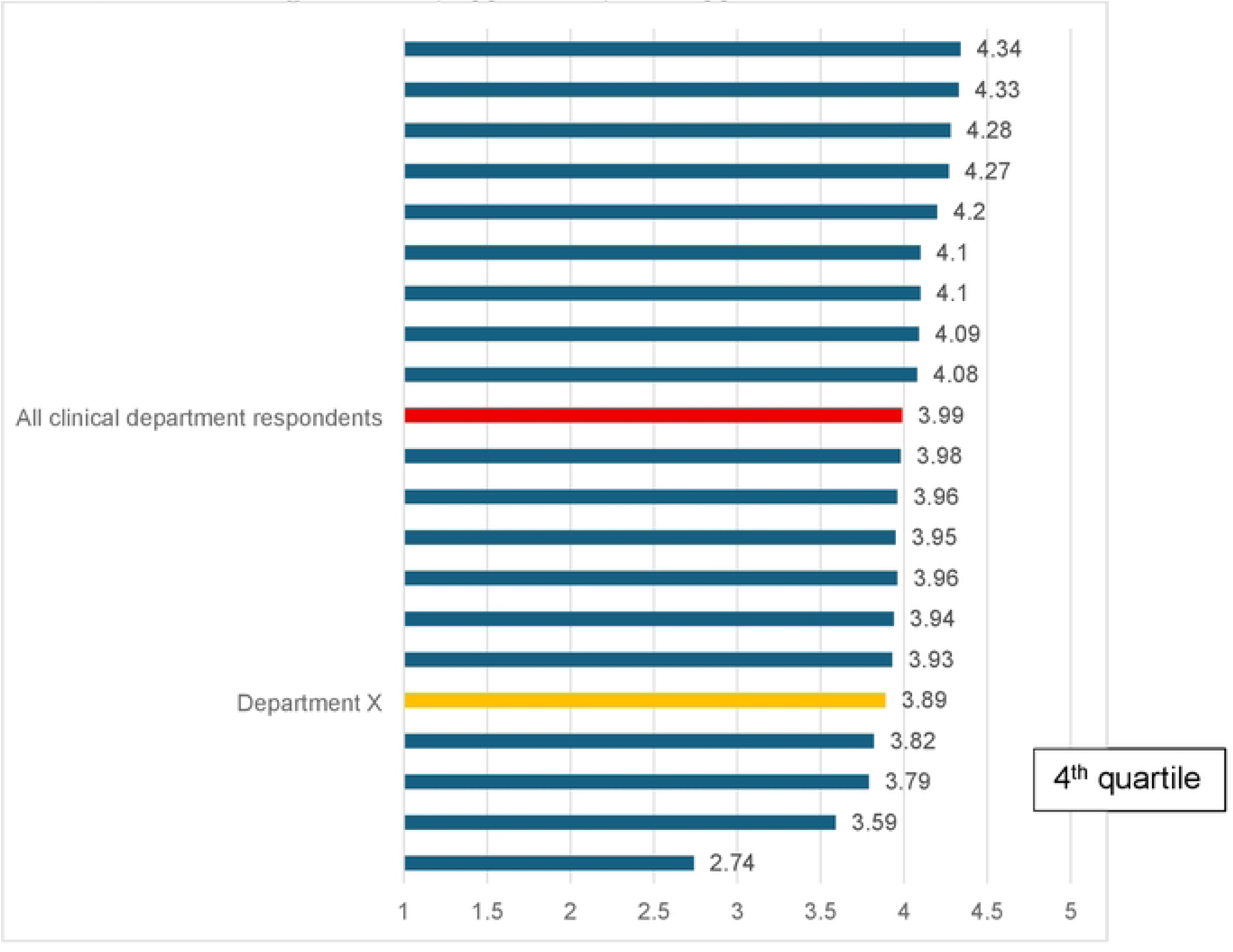
A Department’s Mean Response on an Environment Indicator Arrayed by Mean Responses and Quartile of All Similar Departments. Example of a Clinical Department’s Responses on Dimension 2: Feeling Embraced, Appreciated, and Supported Within the Environment.

**Table 1.** Three Scales of Key Dimensions of the Environment and Their Component Survey Items.

| Dimensions (scale<br>Cronbach's *alpha*) | Component Survey Items |
| --- | --- |
| Perceiving that the unit<br>strives to be supportive (.86) | *Most of my colleagues demonstrate a willingness to understand diversity, equity, and inclusion.* |
|  | *I have noticed efforts to increase diversity in order to reflect the communities we serve.* |
|  | *Diverse backgrounds are taken into consideration when forming teams or committees.* |
|  | *Most of my colleagues are actively engaged in promoting diversity, equity, and inclusion.* |
|  | *Initiatives to promote people's understanding of cultural differences have enhanced relationships among my colleagues.* |
| Feeling embraced,<br>appreciated, and supported<br>within the environment (.94) | *I have someone who supports my success.* |
|  | *I have opportunities for educational and/or professional success that are similar to those of my colleagues.* |
|  | *I have a sense of belonging at [the AHC].* |
|  | *[the AHC] is a place where I am able to perform up to my full potential.* |
|  | *I feel appreciated for my work when it's similar to others who do the same work.* |
|  | *Perspectives like mine are included in decision making.* |
|  | *My accomplishments are recognized similar to others' accomplishments.* |
|  | *All individuals in my role have an equal opportunity for promotion, leadership roles, funding, and compensation.* |
| Feeling accepted in one's beliefs (.79) | *The person(s) I report to/learn from promotes inclusion by valuing individuals based on their diverse backgrounds.* |
|  | *The culture (attitudes, beliefs, and behaviors) where I work or learn aligns with my values.* |
|  | *I can voice an opinion that is different than others without fear of negative consequences.* |

#### Producing individualized department/unit reports

To create individual unit reports, the survey contractor first transferred raw, deidentified individual respondent level survey response data to one faculty member of the AHC survey team. This was the only individual within the AHC to then handle respondent-level data in analyses, to maintain data confidentiality and security.

With more than 50 unique reports needed, including all clinical (19) and basic science (8) departments, large divisions within large departments (6), research and multidisciplinary centers (18), and administrative offices (3), a system was created to standardize and streamline the report generating process. An SPSS program (IBM, Armonk, New York) was created to produce data for each unit with the count and percentage figures required for each graph and table in the order they appeared within reports. A Microsoft Excel report template shell was designed with data tables for each table and figure of the report and prepopulated with data for the appropriate comparison groups (e.g., data for all clinical departments for a report being prepared for a specific clinical department). Research assistants then added information for the specific unit from its SPSS printout into the appropriate tables of the Excel report template. Fully populated Excel Workbooks were then used to generate the tables and graphs then inserted into a Microsoft Word report template, which was prepopulated with graph and table headers and footnotes, standard introductory text, section headers and page breaks. Steps to assure the accuracy of reports included a second research assistant verifying all figures in report drafts against SPSS printouts, and a final review and resolution of any uncertainties by a supervising member of the survey team. Finally, recommendations were added to each report based on that department’s/unit’s areas of strength and weakness in its environment and on the types and sources of mistreatment revealed in the data. Project staff noted each unit’s strengths and weaknesses at each step in the report generating process, with the final choice of specific points, recommendations, and next steps for each report made by the AHC executive leader supporting a healthy environment.

#### Presentations to individual departments/units

Group presentations were held during the late summer and early fall of 2022, scheduled by the chair or director of each of the 50+ departments and other units when their report had been prepared. Attendees included staff, learners and faculty of the unit. The 50 to 60 minute presentations typically occurred at regularly scheduled meetings, such as faculty meetings and grand rounds. Given safety requirements of the COVID-19 pandemic, convenience, and the ability to meet with more people in clinic, lab, or working virtually, presentations were held via Zoom. To convey the importance of the survey and its findings to the unit and to AHC leadership, the unit chair or director introduced the session, and the AHC executive leader supporting a healthy environment presented the report and led the discussion.

Presentations were designed to stimulate interest across subgroups in the environment survey and particularly its findings for their department/unit. Presentations closely followed the flow of information within reports. To start, the survey’s purpose and its goals for departments were briefly reviewed. To build acceptance among attendees that survey findings represented the breadth of views within their department from the people with whom they worked and learned every day, participation rates for the department and for the AHC overall were presented, and the department’s respondents were described in terms of their broad department roles, genders, generations, and other demographics, again to convey the survey’s reach and representativeness.

Having set the stage for attendees, presentations then provided graphic data on how members of the department viewed the environment broadly on the three summary scales. For perspective, mean responses for other similar departments on the scales and overall satisfaction items were presented and the department’s quartile standing among other similar departments on these measures were signposted (Fig 2).

**Figure 2.**
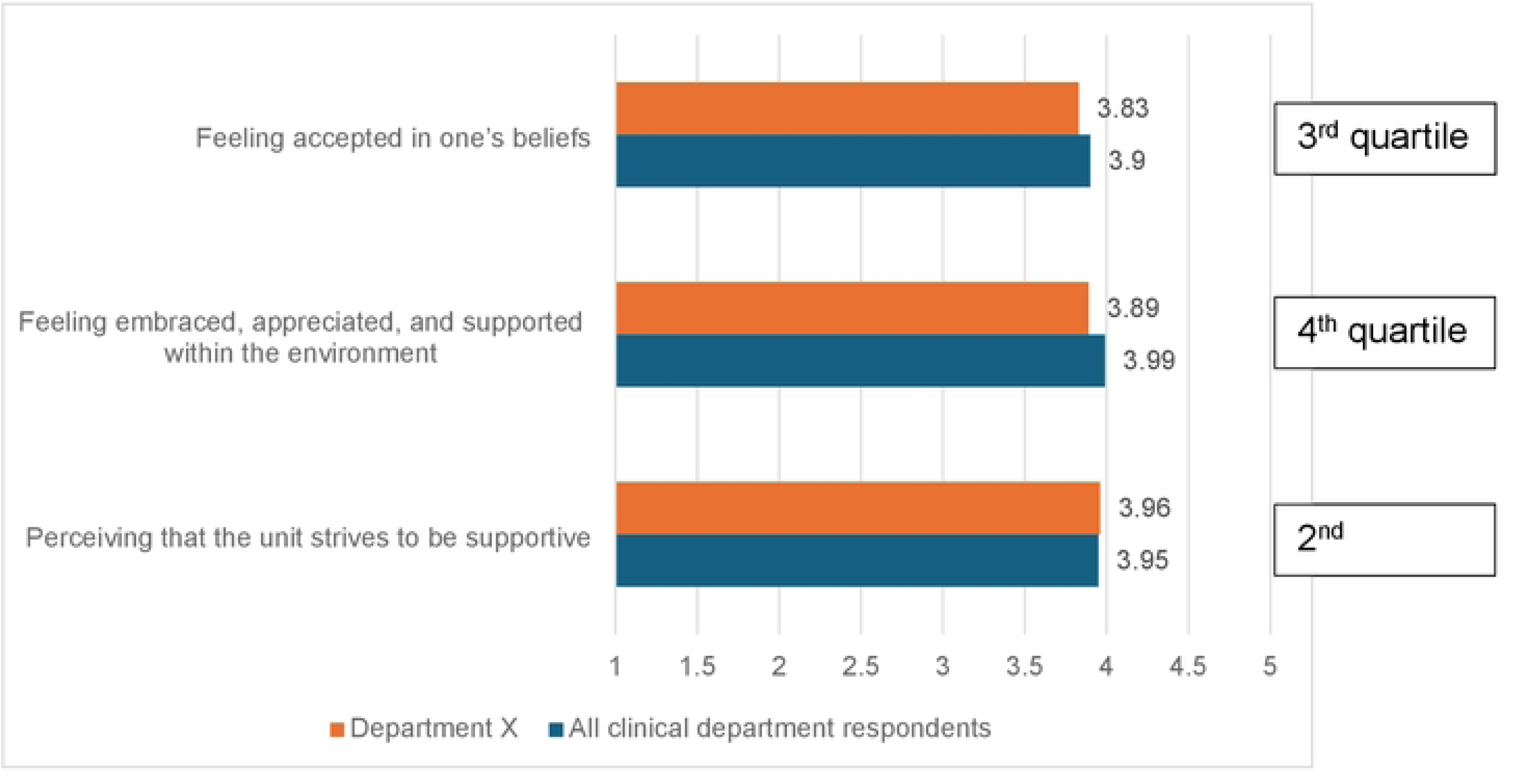
Comparison of Department X and All Other Clinical Departments on the Three Environment Dimension Scales.

Presentations then took attendees on a deeper dive into the data, highlighting first the specific items within each scale on which mean perception ratings within the department differed greatest from that of other departments. Next, the three overall environment scale scores were compared for demographic and department role groups within the department. An initial list of recommendations was then made to address areas where the department’s environment metrics were lowest or were meaningfully lower than similar departments.

Presentations then reviewed survey data on mistreatment, including frequency, knowledge of systems to report and resolve mistreatment, comfort speaking up about mistreatment, rate comparisons across groups within the department and against other departments (Fig 3), and microaggressions (frequency and source) (Fig 4). Recommendations for addressing mistreatment and microaggressions were then provided.

**Figure 3:**
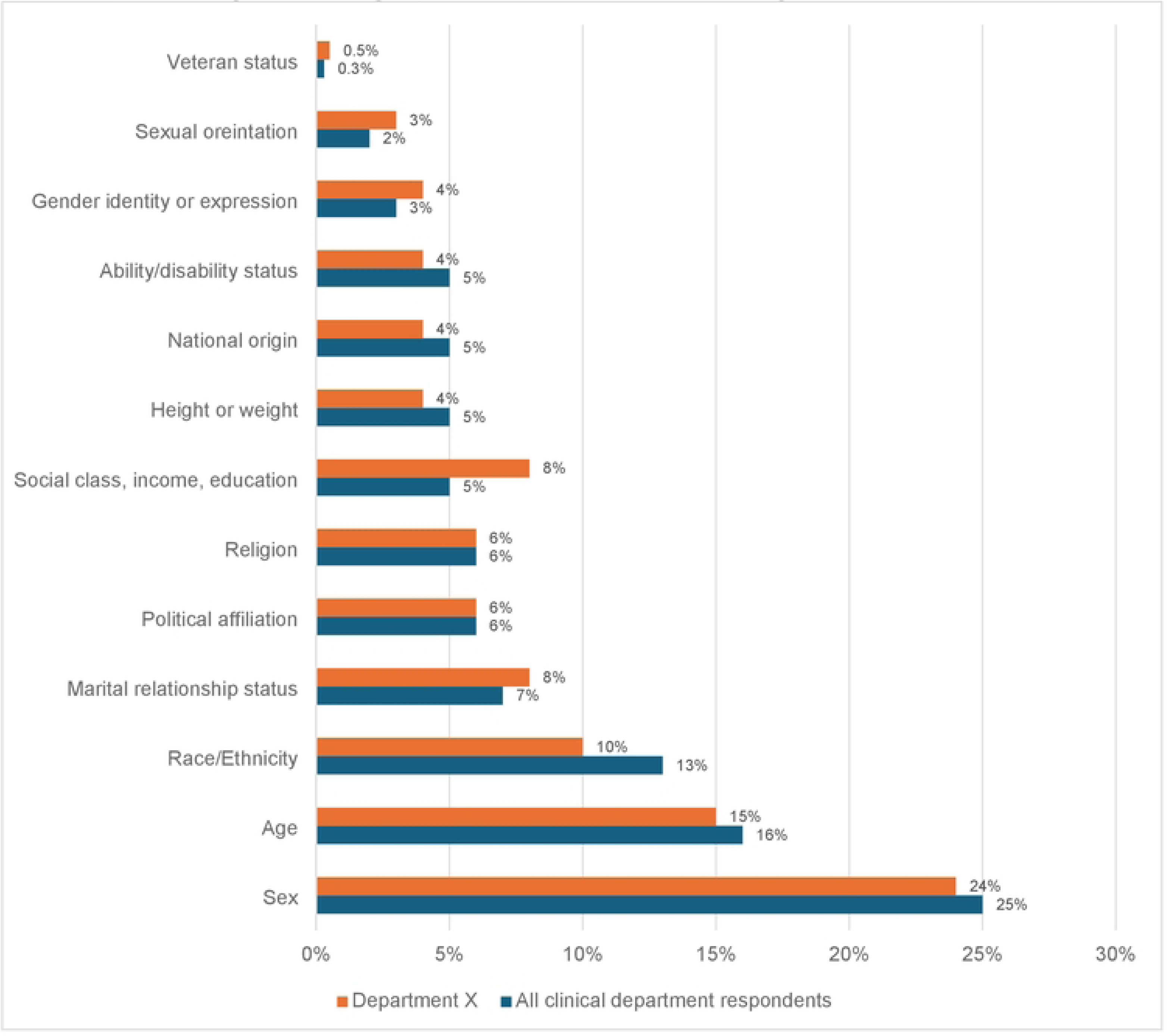
Percent of Respondents Who Personally Experienced Each Type of Mistreatment in the Past 24 Months: Comparison of Department X and All Other Clinical Departments.

**Figure 4:**
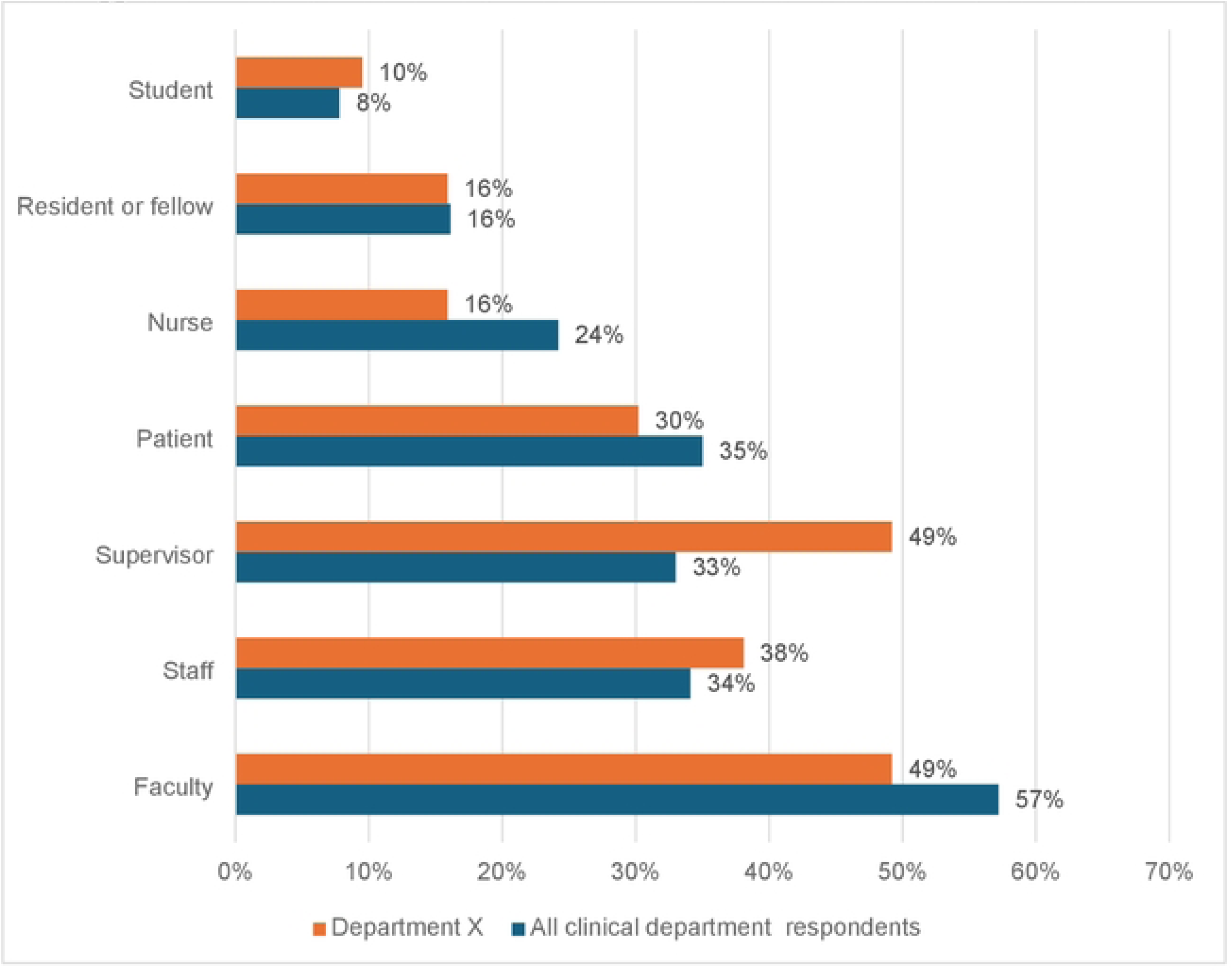
Role of the Offenders Cited by the 63 Department X Respondents Who Experienced Microaggressions: Comparison of Department X and All Other Clinical Departments.

#### Recommendations

The meeting leader then presented recommendations to help departments envision possible next steps. Recommendations included (1) holding large and small group conversations within the department to better understand problem areas identified with low scores, (2) updating existing environment excellence plans to include initiatives addressing newly identified problem areas (3) working with the unit chair or director who can access the AHC’s online interactive summary report to further explore environment strengths and issues across the organization, including for subgroups too small to assess within the department’s own numbers, and (4) addressing low-hanging fruit for some early “wins,” like offering retraining in the AHC processes for reporting mistreatment and developing case studies to augment previous training in recognizing, avoiding and addressing microaggressions and mistreatment. Equally important, attendees were also reminded to celebrate the areas of their department’s demonstrated environment strengths and to build on those strengths. At the end of the presentation, attendees were encouraged to ask questions about the data, its implications for the department, and next steps.

#### Feedback from departments and their subsequent steps

The survey team conducted regular follow-up check-ins with departments’ liaisons to answer any remaining questions about their environment reports and to hear of their departments’ subsequent activities to bolster the environment. In many departments/units, survey findings proved to be catalysts for change, with each using the results in their own ways to inform and stimulate efforts to create a supportive environment.

Departments most often chose to address any low scores on the three broad dimensions of their environment, respond to the action items suggested within reports, and better address microaggressions and mistreatment. Some departments noticed lower scores in units that had graduate students working in research labs or teams. And some departments held workshops and retreats to better understand the lower sense of belonging reported by the unit as a whole or by various subgroups, and to learn how to build a culture of inclusion and belonging through team building, and how to incorporate broad input in administrative decisions. To better address microaggressions and mistreatment, the majority of departments expanded information and education resources (e.g., creating psychologically safe environments) and updated websites for reporting occurrences. Several departments explored expanded financial and administrative support for pathway programs (e.g., rural, low-income, and first-generation learners) to support the next generation of health care professionals. Several departments used newly formed committees and unit-wide meetings to share insights on how to more broadly reduce health disparities through research and clinical practice.

#### Lessons learned

Beyond the lessons noted above at each step of the survey and reporting processes, we note here a few other important lessons learned. Designating and training survey “champions” within each department to coordinate and communicate information about the survey to department members proved critical in building enthusiasm for the survey and gaining strong participation. We do not believe that the alternative, which was to rely on non-prepared administrative staff to promote the survey, would have been as effective.

The amount of time and effort required to create and present reports to 50 individual departments, units, and centers was significant. The five months required for this portion of the process required a combined 15% time of two full-time experienced researchers, substantial time and effort of the AHC’s executive leader for a healthy environment, 50% time of two research assistants, time for department administrative staff to schedule presentations, and one hour from virtually every member of the AHC community to attend their unit’s presentation. This effort resulted in the creation of an online AHC dashboard that resembled the reports for leader access and future data comparison.

Based on our experiences with this survey, our future environment surveys will only ask respondents about their experiences within their departments and not also about their experiences in the broader AHC. During focus groups and at departments’ presentations of survey findings, participants’ comments made it clear that they felt connected to their department, center or learner group, and this was the level they addressed when discussing their environments’ problems and solutions. The AHC-wide environment rarely came up in discussions. However, it is important to ask a few organization-wide questions to inform AHC leadership on mission alignment and a shared organizational culture across all departments and units.

We believe it was the right choice to have enlisted the services of a contractor to help design and conduct the survey. But we learned that the internal survey workgroup had a better understanding of the concepts and measurement of an environment’s supportiveness, and a better appreciation for the importance and effective ways to disseminate environment findings to people in all roles within the AHC and not just leadership. Combining internal and external expertise proved helpful.

## Conclusions

Surveying an AHC community yielded new and actionable information on how the institution’s work and learning environments were perceived and where they could be strengthened. The organization’s understanding of its environment was deepened by assessing people’s experiences in both broad terms—how well they felt embraced and appreciated, felt accepted, and believed the organization strived to be supportive—and through specifics, such as experiences of mistreatment. Analyzing and presenting survey data at the unit level – e.g., individual department, center, office, and training program level - matched people’s notions of what constituted “their environment.” Information presented at this level and to all members of each unit, therefore, garnered wide interest and helped organizational members become more informed on, sensitive to, and subsequently more open to talking about the environment explicitly, along with potential and necessary directions for change.

While being mindful of lessons we learned, the survey and dissemination steps we undertook should be appropriate for other AHC’s wanting to create an organizational culture that better understand the strengths and weakness of their environments in the eyes of the people who work and learn there. Important requirements for such a survey include adequate time and funding, local and contracted expertise, and supportive leadership. Individual AHCs may want to further expand, amend or refocus some of the questions asked in questionnaires, as we believe that environment surveys will appropriately continue to evolve and strengthen.

AHCs may be best advised to focus surveys on the broad and less contentious aspects of the environment, like people’s sense of being appreciated and supported. Similarly, for now, equity issues in the environment could be limited to comparisons of individuals across role groups, geographic location (rural vs. non-rural or in state vs. out of state), generations, education-level groups, and tenured vs. non-tenured faculty, and take a deeper dive in understanding perspectives in the AHC.

## Data Availability

Data for this study are HR protected and cannot be shared publicly. This study does not focus on data/results, but process and approach. Therefore, data sharing would not be appropriate.

## Acknowledgements

The authors appreciate the many individuals across the institution for their invaluable help creating and implementing the environment survey, encouraging survey participation, and organizing presentations of findings to departments. The authors specifically thank Aleyah Pankey, Ingrid Jones, Victor Reese, Sam Hofstetter, Mark Poklar, and Virginia Nadworny.

## Notes

### Competing Interest Statement

The authors have declared no competing interest.

### Funding Statement

The author(s) received no specific funding for this work.

### Author Declarations

This submission, Reference ID 339715, was reviewed by the Office of Human Research Ethics at the University of North Carolina at Chapel Hill, which has determined that this submission does not constitute human subjects research as defined under federal regulations [45 CFR 46.102 (e or l) and 21 CFR 56.102(c)(e)(l)] and does not require IRB approval.

